# Age significantly influences the sensitivity of SARS-CoV-2 rapid antibody assays

**DOI:** 10.1101/2021.01.28.21250675

**Authors:** Natalie Irwin, Lyle Murray, Benjamin Ozynski, Guy A Richards, Graham Paget, Jacqueline Venturas, Ismail Kalla, Nina Diana, Adam Mahomed, Jarrod Zamparini

**Affiliations:** Department of Internal Medicine, Faculty of Health Sciences, University of the Witwatersrand, Johannesburg, South Africa; Department of Medicine, Charlotte Maxeke Johannesburg Academic Hospital, Johannesburg, South Africa; Wits Health Innovation, Johannesburg, South Africa

## Abstract

**BACKGROUND:** Point of care serological assays are a promising tool in COVID-19 diagnostics but do have limitations. This study evaluated the sensitivity of five rapid antibody assays and explored factors influencing their sensitivity to detect SARS-CoV-2-specific IgG and IgM antibodies.

**METHODS:** Finger-prick blood samples from 102 participants, within two to six weeks of PCR-confirmed COVID-19 diagnosis, were tested for IgG and IgM on five rapid serological assays. The assay sensitivities were compared, and patient factors evaluated in order to investigate potential associations with assay sensitivity.

**RESULTS:** Sensitivity ranged from 36% to 69% for IgG and 13% to 67% for IgM. Age was the only factor significantly influencing the likelihood of a detectable IgG or IgM response. Individuals aged 40 years and older had an increased likelihood of a detectable IgG or IgM antibody response by rapid antibody assay.

**CONCLUSION:** Rapid serological assays demonstrate significant variability when used in a real-world clinical context. There may be limitations in their use for COVID-19 diagnosis amongst the young.

## BACKGROUND

Rapid serological assays, used at the point of care (POC), pose a promising clinical tool in the diagnosis of Coronavirus disease 2019 (COVID-19), particularly in low- and middle-income countries where diagnostic resources are scarce. These lateral flow chromatographic immunoassays qualitatively detect immunoglobulin G (IgG) and immunoglobulin M (IgM) antibodies to the severe acute respiratory syndrome coronavirus 2 (SARS-CoV-2) on a venous or finger-prick whole blood sample without the need for specialized equipment. Such assays are useful for rapid antibody testing in surveillance programmes in outbreak settings or in high seroprevalence areas. The assays may assist in the diagnosis of suspected COVID-19 in patients who test negative for SARS-CoV-2 by polymerase chain reaction (PCR) on naso- or oropharyngeal swabs.[1] In addition, they require minimal operator training and have a turnaround time of under thirty minutes.[2]

Some valid concerns about the performance quality of these rapid assays exist, and most available rapid assays have been subjected only to single centre internal validation studies, in small populations.[3] Furthermore, the threshold antibody titre required to generate a detectable result on these devices is poorly described. Reported overall IgG/IgM sensitivities range from 18.4 to 93.3% and vary depending on disease severity and duration since symptom onset.[2]

This study critically evaluated the sensitivity of five rapid antibody assays for detection of SARS-CoV-2-specific IgM and IgG antibodies, on finger-prick blood samples amongst patients with COVID-19 confirmed by PCR on nasopharyngeal or oropharyngeal swab. Importantly, this study also investigates patient factors that influence the sensitivity of such assays.

## METHODS AND MATERIALS

This study was approved by the University of the Witwatersrand Human Research Ethics Committee (Medical) (M200697). Written informed consent was obtained from all participants and patient data were anonymised prior to analysis.

### Study participants

Adult participants (≥18 years old) were recruited at the Charlotte Maxeke Johannesburg Academic Hospital in Johannesburg, South Africa. Randomly selected inpatients and outpatients were invited to participate if they had laboratory confirmed COVID-19 by RT-PCR on a naso- or oropharyngeal swab prior to interview and testing. Participant numbers were restricted by the number of assay cassettes available.

Clinical and biographical data were collected using an electronic database (REDCap 10.6.2, Vanderbilt University) by means of a self-administered participant questionnaire. Variables collected included demographics (age, sex, self-reported ethnicity), co-morbidities and if the participant was a healthcare worker (HCW). Participants provided details of previous PCR testing including the number of previous tests done, the date and result of each test, the route of sampling (oropharyngeal or nasopharyngeal), symptoms experienced at the time of the positive test, the date of onset of symptoms and the severity of disease (asymptomatic, mild, moderate, severe, critical) classified according to published criteria.[4]

### Rapid Antibody Assays

Five rapid immunochromatographic antibody assays were evaluated in this study and performed for each participant, namely:

1. 2019-nCoV-IgG/IgM Rapid Test (whole blood, serum or plasma), Lot 200505, Dynamiker Biotechnology Company Ltd, Tianjin, China (Dynamiker)
2. 2019-nCoV IgG/IgM Rapid Test Cassette (whole blood, serum or plasma), Lot NCP20030123, AllTest Biotech Company Ltd, Hangzhou, China (AllTest)
3. 2019-nCoV Ab Test (Colloidal Gold) (serum, plasma, venous whole blood), Lot 20200402, Innovita Biotechnology Company Ltd, Tangshan, China (Innovita)
4. Medical Diagnostech COVID-19 IgG/IgM Rapid test (whole blood, serum or plasma), Lot 200703, Altis Biologics (Pty) Ltd, Pretoria, South Africa (Altis)
5. Cellex qSARS-CoV-2 IgG/IgM Cassette Rapid Test (serum, plasma, whole blood), Lot WI1106C-DH-GZ-20200511, Cellex, Jiangsu, China (Cellex)

A single drop (10-20μL) of whole blood drawn by fingertip puncture was deposited in the sample well of each test cassette. Two to five drops of reagent buffer were then added to the sample well and results read 15-20 minutes later according to the specific manufacturer’s instructions. Assays three and five had not previously been validated on finger-prick specimens. Two readers (N.I. and/or J.Z. and/or B.O.) read the cassettes with the naked eye and a third reader (J.V.) settled any disputes. A test was considered valid if a control line was visualized and was interpreted as positive if the control line and the line for IgG or IgM or both was seen.

### Statistical analysis

Data were analysed using Prism 8.4.3 (GraphPad Software Inc, La Jolla, California) using standard non-parametric statistical tests as appropriate. Continuous data were expressed as medians with interquartile ranges (IQRs) and categorical variables presented as numbers and percentages. Fisher’s exact tests were used to compare results in the age, time and severity groups and Spearman’s correlation co-efficients were used to assess agreement between test assays. The multivariate logistic regression analysis was performed using IBM SPSS Statistics 26.0 (IBM Corporation, Armonk, New York).

## RESULTS

### Demographics

We conducted antibody testing using all five rapid antibody assays on 102 participants with previous PCR-confirmed COVID-19. The majority of our participants were female (61%) and of black ethnicity (40%). Median age in the cohort was 37.5 years (IQR 29 – 45.75) and most of the participants were HCWs (68%). The majority of our cohort were tested between 15- and 42-days post-positive PCR testing (n=83, 81%) with 4 (4%) and 15 (15%) being tested less than 15 days and more than 42 days post-testing, respectively. Comorbidities amongst the participants included hypertension (n=14, 14%), diabetes (n=12, 12%), asthma (n=10, 10%), HIV (n=6, 6%) and cancer (n=3, 3%). Additional demographic data are presented in Table 1.

**Table 1.**
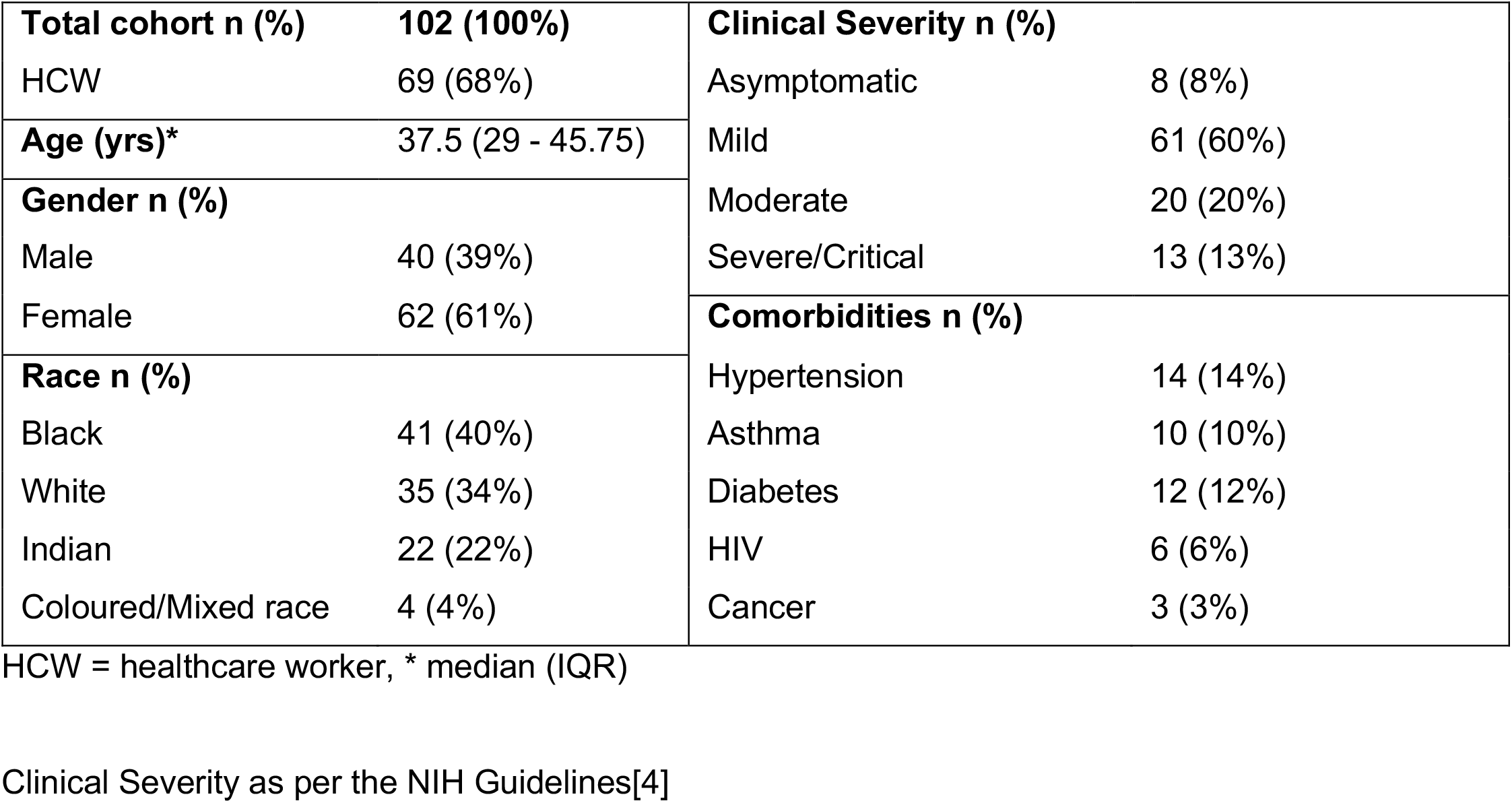
Participant Demographics

|  |  |  |  |
| --- | --- | --- | --- |
| **Total cohort n (%)** | **102 (100%)** | **Clinical Severity n (%)** |  |
| HCW | 69 (68%) | Asymptomatic | 8 (8%) |
| **Age (yrs)\*** | **37.5 (29 - 45.75)** | Mild | 61 (60%) |
| **Gender n (%)** |  | Moderate | 20 (20%) |
| Male | 40 (39%) | Severe/Critical | 13 (13%) |
| Female | 62 (61%) | **Comorbidities n (%)** |  |
| **Race n (%)** |  | Hypertension | 14 (14%) |
| Black | 41 (40%) | Asthma | 10 (10%) |
| White | 35 (34%) | Diabetes | 12 (12%) |
| Indian | 22 (22%) | HIV | 6 (6%) |
| Coloured/Mixed race | 4 (4%) | Cancer | 3 (3%) |

HCW = healthcare worker, \* median (IQR)
Clinical Severity as per the NIH Guidelines[4]

### Antibody assay sensitivity

Overall sensitivity to detect SARS-CoV-2-specific IgG and IgM antibodies was below 70% in all assays (Figure 1). IgG sensitivity ranged from 36% (Innovita) to 69% (Dynamiker) whilst IgM sensitivity ranged from 13% (Innovita) to 67% (Dynamiker). Of note, the sensitivities of the Innovita (13%), AllTest (15%) and Altis (26%) assays to detect IgM were markedly lower than those of the Dynamiker (67%) and Cellex (64%) assays.

**Figure 1.**
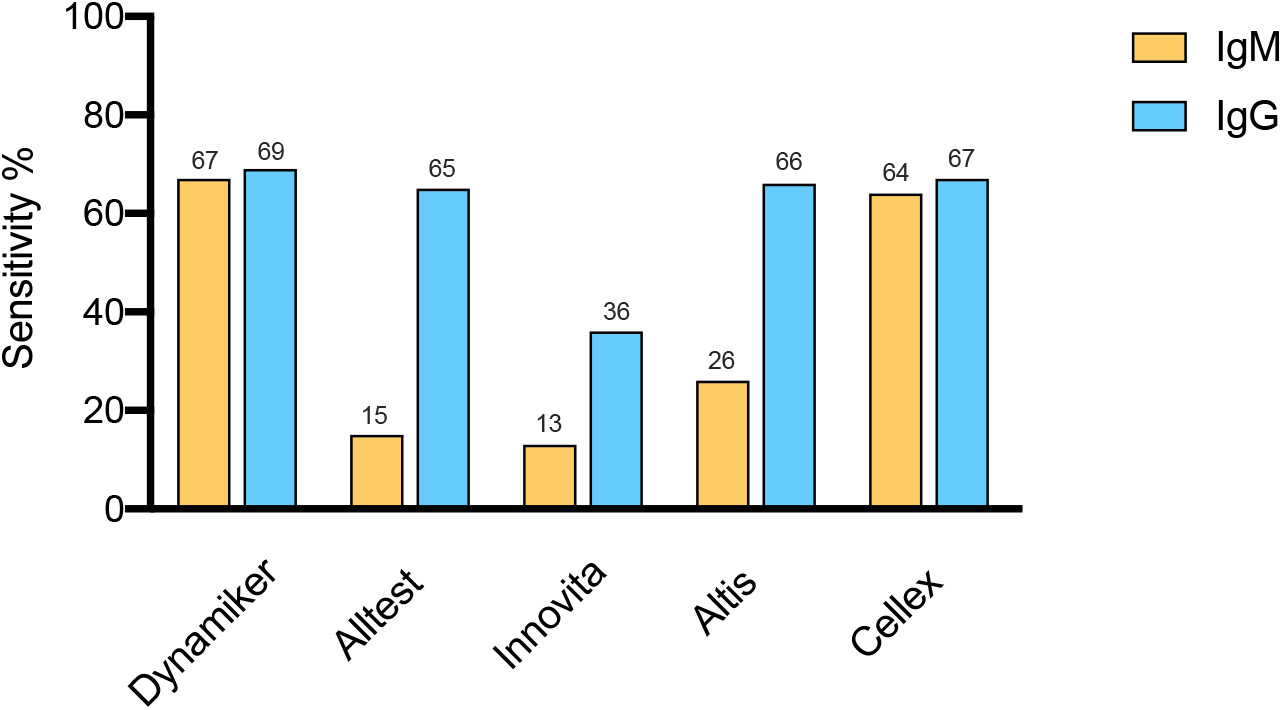
Total sensitivity of five rapid antibody assays. Bar graphs show the sensitivity of each rapid antibody assay in detecting SARS-CoV-2-specific IgG and IgM in the total cohort (n=102). The values above each bar are percentages.

### Variability between assays

Variability in diagnostic accuracy between the five rapid antibody assays for the detection IgG and/or IgM was assessed through the use of a correlation matrix represented in a heatmap (Figure 2). This measured agreement between each assay for the detection of IgG or IgM. Four of the five assays (Dynamiker, AllTest, Altis and Cellex, Figure 2; r >0.8 for all) correlated well with each other in the detection of IgG. However, none of these four assays correlated well with the Innovita assay for IgG. Strong correlation for IgM results was found between only two of the five assays (Dynamiker and Cellex, Figure 2; r >0.8). The correlation results were all statistically significant at p<0.05.

**Figure 2.**
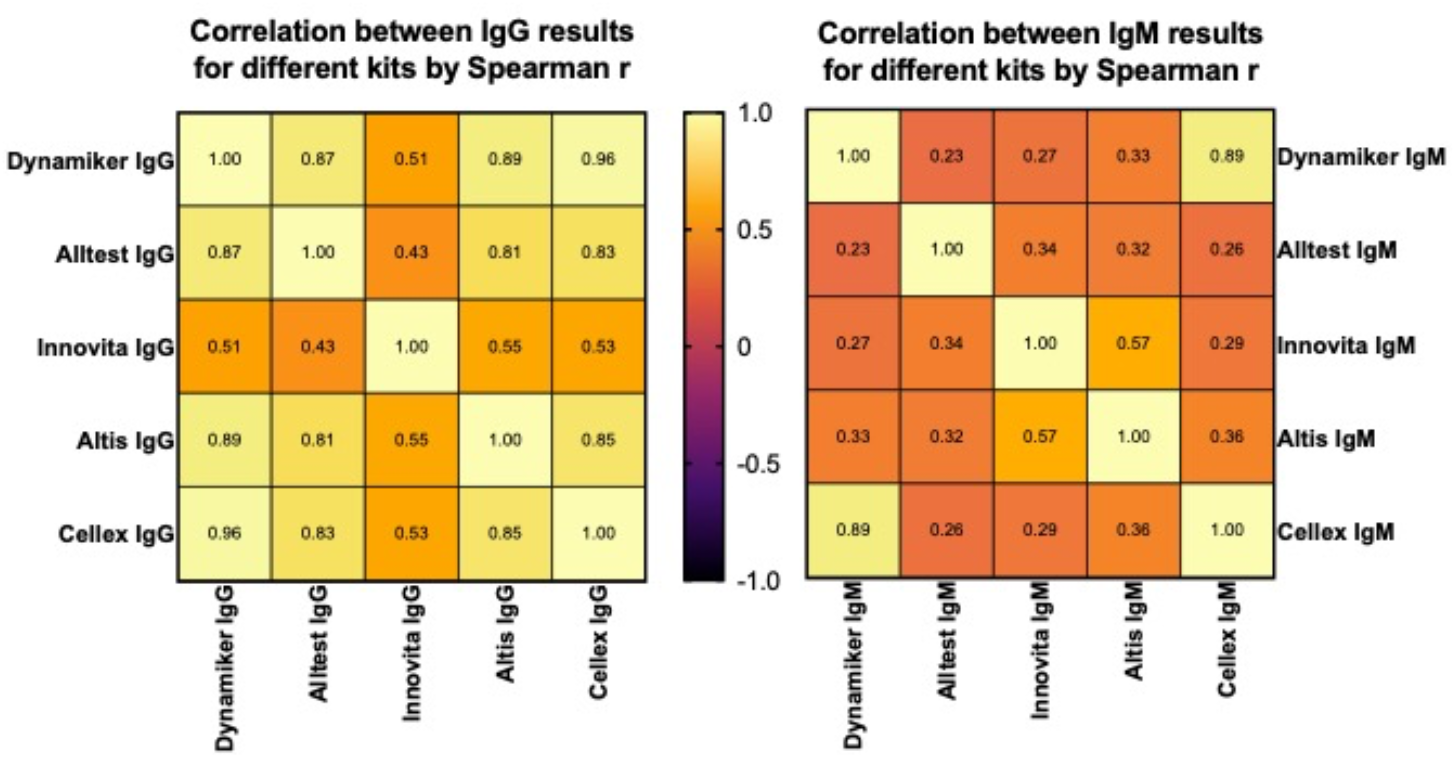
Heat map showing the correlation between each rapid antibody assay for the detection of SARS-CoV-2-specific IgG and IgM. The Spearman “r” coefficient showing the degree of agreement between each pair of rapid antibody assays is shown in each block for IgG (left panel) and IgM (right panel). A Spearman “r” value of greater than 0.8 is considered significant agreement. All values shown had significant P values of <0.05

### Analysis of the factors associated with improved sensitivity of the rapid antibody assays

The participant factors potentially associated with the sensitivity for detection of IgG or IgM by rapid antibody assay were investigated and included: gender, time since positive SARS-CoV-2 PCR test (≤30 days vs >30 days), age (<40 years vs ≥40 years) and COVID-19 disease severity (asymptomatic-mild vs moderate-severe).

#### Age

Most strikingly, there was a significant reduction in IgG sensitivity for participants under 40 years of age compared to those over 40 years for all of the assays used (Table 2, p<0.005 for all). Similarly there was a significant reduction in IgM sensitivity in those under 40 years of age compared to those over 40 years of age, however, this significance was only demonstrated in four of the five assays (Table 3, p<0.005 for all except the AllTest).

**Table 2.** Participant factors associated with IgG sensitivity

|  | Dynamiker IgG | AllTest IgG | Innovita IgG | Altis IgG | Cellex IgG |
| --- | --- | --- | --- | --- | --- |
| **Age** |  |  |  |  |  |
| <40 years (n=56) | 29 (52%) | 28 (50%) | 11 (20%) | 28 (50%) | 28 (50%) |
| ≥40 years (n=46) | 41 (89%) | 38 (83%) | 26 (57%) | 39 (85%) | 40 (87%) |
| P (Fisher's exact) | <0.0001 | 0.0008 | 0.0002 | 0.0003 | 0.0001 |
| OR (95%CI) | 7.6 (2.6 - 19.6) | 4.8 (1.9 - 11.8) | 5.3 (2.3 - 12.9) | 5.6 (2.2 - 13.7) | 6.7 (2. - 18.0) |
| **Gender** |  |  |  |  |  |
| Male (n=40) | 25 (63%) | 24 (60%) | 15 (38%) | 22 (55%) | 25 (63%) |
| Female (n=62) | 45 (73%) | 42 (68%) | 22 (36%) | 45 (73%) | 43 (69%) |
| P (Fisher's exact) | 0.3822 | 0.525 | 0.8365 | 0.0881 | 0.5226 |
| (95%CI) | 0.6 (0.3 – 1.5) | 0.7 (0.3 – 1.6) | 1.1 (0.5 – 2.4) | 0.5 (0.2 – 1.0) | 0.7 (0.3 – 1.7) |
| **Time since positive PCR** |  |  |  |  |  |
| ≤30 days (n=53) | 33 (62%) | 31 (58%) | 20 (38%) | 32 (60%) | 33 (62%) |
| >30 days (n=49) | 37 (76%) | 35 (71%) | 17 (35%) | 35 (71%) | 35 (71%) |
| P (Fisher's exact) | 0.2004 | 0.2149 | 0.8376 | 0.2982 | 0.4019 |
| (95%CI) | 1.9 (0.8 - 4.2) | 1.8 (0.7 - 4.1) | 0.9 (0.4 - 1.9) | 1.6 (0.7 - 3.8) | 1.5 (0.7 - 3.6) |
| **Disease severity** |  |  |  |  |  |
| Asymptomatic/Mild (n=69) | 45 (65%) | 42 (61%) | 21 (30%) | 43 (62%) | 44 (64%) |
| Moderate/Severe (n=33) | 25 (76%) | 24 (73%) | 16 (48%) | 24 (73%) | 24 (73%) |
| P (Fisher's exact) | 0.3636 | 0.2749 | 0.0837 | 0.375 | 0.5011 |
| (95%CI) | 1.7 (0.6 - 4.2) | 1.7 (0.7 - 4.0) | 2.1 (0.9 - 4.9) | 1.6 (0.7 - 3.8) | 1.5 (0.6 - 3.5) |

OR=Odds Ratio, 95%CI = 95% Confidence Interval.

**Table 3.** Participant factors associated with IgM sensitivity

|  | Dynamiker IgM | AllTest IgM | Innovita IgM | Altis IgM | Cellex IgM |
| --- | --- | --- | --- | --- | --- |
| **Age** |  |  |  |  |  |
| <40 years (n=56) | 29 (52%) | 5 (9%) | 2 (4%) | 8 (14%) | 25 (45%) |
| ≥40 years (n=46) | 39 (85%) | 10 (22%) | 11 (24%) | 19 (41%) | 40 (87%) |
| P (Fisher's exact) | 0.0006 | 0.093 | 0.0026 | 0.0031 | <0.0001 |
| (95%CI) | 5.2 (2.0 - 12.8) | 2.8 (0.9 - 7.9) | 8.5 (1.8 - 39.5) | 4.2 (1.6 - 10.9) | 8.3 (3.0 - 22.3) |
| **Gender** |  |  |  |  |  |
| Male (n=40) | 25 (63%) | 6 (15%) | 7 (18%) | 14 (35%) | 23 (58%) |
| Female (n=62) | 43 (69%) | 9 (15%) | 6 (10%) | 13 (21%) | 42 (68%) |
| P (Fisher's exact) | 0.5226 | 0.99 | 0.3621 | 0.1673 | 0.3018 |
| (95%CI) | 0.7 (0.3 - 1.7) | 1.0 (0.3 – 3.2) | 2.0 (0.7 – 6.8) | 2.0 (0.8 – 4.8) | 0.6 (0.3 – 1.4) |
| **Time since positive PCR** |  |  |  |  |  |
| ≤30 days (n=53) | 32 (60%) | 12 (23%) | 7 (13%) | 16 (30%) | 33 (62%) |
| >30 days (n=49) | 36 (73%) | 3 (6%) | 6 (12%) | 11 (22%) | 32 (65%) |
| P (Fisher's exact) | 0.2081 | 0.0246 | 1.0 | 0.5010 | 0.8376 |
| (95%CI) | 1.8 (0.8 - 4.2) | 0.2 (0.1 – 0.8) | 0.9 (0.3 - 2.8) | 0.7 (0.3 - 1.7) | 1.1 (0.5 - 2.5) |
| **Disease severity** |  |  |  |  |  |
| Asymptomatic/Mild (n=69) | 44 (64%) | 9 (13%) | 7 (10%) | 15 (22%) | 41 (59%) |
| Moderate/Severe (n=33) | 24 (73%) | 6 (18%) | 6 (18%) | 12 (36%) | 24 (73%) |
| P (Fisher's exact) | 0.5011 | 0.5547 | 0.3415 | 0.1509 | 0.2711 |
| (95%CI) | 1.5 (0.6 - 3.5) | 1.5 (0.5 - 4.6) | 2.0 (0.6 - 6.0) | 2.1 (0.8 - 4.9) | 1.8 (0.7 - 4.2) |

OR=Odds Ratio, 95%CI = 95% Confidence Interval.

#### Gender

Male gender has previously been associated with COVID-19 severity.[5] However, we did not find any difference in the sensitivity of any of the rapid antibody assays for IgG or IgM based on gender within our cohort (Tables 2 and 3).

#### Time since positive SARS-CoV-2 PCR test

We hypothesized that the dynamic antibody responses during and after COVID-19 may influence the detection of IgG and IgM SARS-CoV-2-specific antibodies. The association between time since the SARS-CoV-2 PCR test and the detection of IgG and IgM antibodies was explored. There was no difference in IgG sensitivity between those presenting within 30 days of a positive PCR test compared to those presenting after 30 days as shown in Table 2. However, a significantly lower proportion of individuals tested more than 30 days after a positive SARS-CoV-2 PCR test had a detectable IgM response by the AllTest assay (23% (≤30 days) vs 6% (>30 days), p = 0.02, OR 0.2228(0.0644 - 0.7993), Table 3). There was no significant difference in IgM sensitivity in the two time groups for the other four rapid antibody assays (p>0.05 for Dynamiker, Innovita, Altis and Cellex, Table 3).

#### Disease severity

COVID-19 disease severity has been shown to impact the magnitude of the SARS-CoV-2-specific antibody response.[6] We therefore investigated whether disease severity had an influence on the sensitivities of the rapid antibody assays to detect IgG or IgM antibodies to SARS-CoV-2. Interestingly, there was no significant difference in IgG sensitivity for those with asymptomatic or mild disease compared to those with moderate or severe disease between the two groups for each assay individually (Table 2). There was also no significant difference in IgM sensitivity between the two groups (p>0.05 for all, Table 3)

#### Multivariate analysis

A multivariate logistic regression was performed to exclude confounding variables and to confirm the factors associated with an increased likelihood of a detectable SARS-CoV-2-specific IgG or IgM response by rapid antibody assay. The variables included age, gender, time since positive test and severity. Age >40 years was the only variable associated with a significantly increased likelihood of a detectable IgG and IgM response by rapid antibody testing (Supplementary Tables 1 and 2). This was significant for all of the rapid antibody assays for IgG and all except the AllTest assay for IgM (Supplementary Tables 1 and 2).

## DISCUSSION

Rapid serological assays are increasingly becoming an essential component of surveillance of outbreaks and retrospective diagnoses of COVID-19. These assays have appeal as they are rapid, inexpensive and user-friendly. However, data evaluating their true clinical performance on finger-prick analyses at the POC, in a real-world clinical context, is urgently needed.

In this present study, tested against the reference gold standard PCR, the sensitivity of five rapid antibody assays ranged from 36% to 69% for IgG and 13% to 67% for IgM. This is significantly lower than that reported in previous validation studies of these rapid tests.[7–9] The South African Health Products Regulatory Authority (SAHPRA) specifies that a minimum clinical sensitivity of 85% within 95% confidence intervals be proven prior to registration.[10] All of these rapid antibody assays fall below that benchmark when used in our cohort. The wide spread of the results of our sensitivity analysis are comparable with findings of existing studies. In a pooled analysis of 1030 POC antibody assays, by Riccò *et al*, combined sensitivity for IgG and IgM, ranged from 18.4% to 93.3% and with an average of 64.8% (95%CI 54.5–74.0).[2] We also report poor correlation between the results of the five assays in comparison to each other, particularly when testing for IgM. This finding has also been previously described, and prompted Van Elslande *et al* to question the need for measuring IgM SARS-CoV-2 antibodies at all.[11,12] The heterogenous clinical sensitivity performance of IgM in the antibody assays in our study further suggest limited clinical utility.

Our analysis of patient factors associated with an improved sensitivity of the rapid antibody tests revealed an association between age and sensitivity. All of the assays demonstrated improved sensitivities in those patients aged >40 years for IgG and all the assays except the Altis for IgM. The higher sensitivity of these tests in older participants may indicate higher antibody titres in these individuals and therefore a higher likelihood of detection of such antibody responses by the rapid assays. Older patients with COVID-19 have been shown to have higher SARS-CoV-2-specific IgG and IgM antibody titres than younger patients, although the reason for this is unclear.[13,14] It is likely that increased SARS-CoV-2-specific antibody titres correlate with more severe COVID-19, and that increased age has been correlated with severe disease and worse outcome.[6,15]. When both age and disease severity were included in a multivariate logistic regression analysis, only age was associated with an increased likelihood of a positive rapid IgG or IgM antibody response, thereby suggesting that age may play a role independently of its association with disease severity. Our cohort primarily consisted of participants that had mild COVID-19 with 80% of the cohort in the mild to moderate category. Due to the fact that younger individuals typically experience milder COVID-19 disease and have an increased rate of asymptomatic infection, our findings may suggest a significant impediment to the use of these assays to assess seroprevalence in younger participants.

There are limitations to this study. Although used as the reference gold standard, PCR-based testing gives no indication of patient seropositivity. The negative results reported may represent low antibody titres or low participant seroconversion, rather than a failure of the test to detect antibodies. Furthermore, two assays (Innovita and Cellex) that were included have not previously been validated on finger-prick blood samples. Although we consider the effect is likely to be small, as all other tests could be used across blood sample platforms, this may have contributed to their poor performance. Owing to inadequate access to negative control samples, we were not able to perform a corresponding specificity analysis.

## CONCLUSION

This study described an overall underperformance of rapid serological assays to detect an IgG and IgM response two to six weeks after PCR confirmed SARS-CoV-2 infection. We highlight the heterogenous ability of the antibody assays to detect IgM and describe a significant independent association between age >40 years and increased sensitivity for IgG and IgM seropositivity. Judicious clinical use, recognising the limitations of rapid serological assays, especially amongst the young, is necessary.

## Data Availability

All data is available from Dr Natalie Irwin and/or Dr Jarrod Zamparini

## ACKNOWLEDGEMENTS

We would like to acknowledge First Medical Company, SMD Technologies and Altis Biologics for donating test assays for use in this study.

