## Supplemental Tables for "Age significantly influences the sensitivity of SARS-CoV-2 rapid antibody assays"

Supplemental Data

IgG Multivariate Data

| Variable | Dynamiker IgM | | AllTest IgM | | Innovita IgM | | Altis IgM | | Cellex IgM | |
|---|---|---|---|---|---|---|---|---|---|---|
| | *OR (95%CI)* | *P* | *OR (95%CI)* | *P* | *OR (95%CI)* | *P* | *OR (95%CI)* | *P* | *OR (95%CI)* | *P* |
| **Age** | | | | | | | | | | |
| - **<40 years** | 1 | | 1 | | 1 | | 1 | | 1 | |
| - **>40 years** | 7.8 (2.6 - 23.6) | **<0.001** | 4.6 (1.8 – 12.1) | **0.002** | 4.9 (2.0 – 12.2) | **0.001** | 5.7 (2.1 – 15.6) | **0.001** | 6.8 (2.4 – 19.1) | **<0.001** |
| **Gender** | | | | | | | | | | |
| - **Male** | 1 | | 1 | | 1 | | 1 | | 1 | |
| - **Female** | 0.6 (0.3 – 1.7) | 0.36 | 0.8 (0.3 – 1.8) | 0.53 | 1.2 (0.50 – 2.9) | 0.76 | 0.50 (0.2 – 1.1) | 0.09 | 0.8 (0.3 – 1.9) | 0.59 |
| **Disease severity** | | | | | | | | | | |
| - **Asym - mild** | 1 | | 1 | | 1 | | 1 | | 1 | |
| - **Mod - Sev** | 1.3 (0.4 – 3.8) | 0.69 | 1.4 (0.5 – 3.8) | 0.53 | 1.5 (0.6 – 3.8) | 0.42 | 1.2 (0.4 – 3.5) | 0.70 | 1.1 (0.4 – 3.0) | 0.93 |
| **Time since positive PCR** | | | | | | | | | | |
| - **<30 days** | 1 | | 1 | | 1 | | 1 | | 1 | |
| - **>30 days** | 2.2 (0.8 – 5.8) | 0.11 | 2.0 (0.8 – 5.0) | 0.13 | 0.9 (0.40 – 2.2) | 0.82 | 1.8 (0.7 – 4.6) | 0.22 | 1.6 (0.70 – 4.2) | 0.30 |

Irwin N, Murray L, Ozynski B, Richards GA, Paget G, Venturas J, Kalla I, Diana N, Mahomed A, Zamparini J. **Age significantly influences the sensitivity of SARS-CoV-2 rapid antibody assays**

IgM Multivariate Data

| Variable | Dynamiker IgM | | AllTest IgM | | Innovita IgM | | Altis IgM | | Cellex IgM | |
|---|---|---|---|---|---|---|---|---|---|---|
| | *OR (95%CI)* | *P* | *OR (95%CI)* | *P* | *OR (95%CI)* | *P* | *OR (95%CI)* | *P* | *OR (95%CI)* | *P* |
| **Age** | | | | | | | | | | |
| - **<40 yrs** | 1 | | 1 | | 1 | | 1 | | 1 | |
| - **>40 yrs** | 5.2 (1.9 – 14.2) | **0.001** | 3.0 (0.9 – 9.9) | 0.09 | 8.9 (1.8 – 45.4) | **0.009** | 4.3 (1.6 – 11.7) | **0.005** | 8.0 (2.8 – 22.7) | **<0.001** |
| **Gender** | | | | | | | | | | |
| - **Male** | 1 | | 1 | | 1 | | 1 | | 1 | |
| - **Female** | 0.8 (0.3 – 1.9) | 0.60 | 0.9 (0.3 – 3.0) | 0.86 | 2.3 (0.7 – 8.4) | 0.20 | 2.2 (0.9 – 5.8) | 0.11 | 0.6 (0.3 – 1.6) | 0.35 |
| **Disease severity** | | | | | | | | | | |
| - **Asym - mild** | 1 | | 1 | | 1 | | 1 | | 1 | |
| - **Mod - Sev** | 1.2 (0.4 – 3.3) | 0.77 | 1.1 (0.3 – 3.6) | 0.89 | 1.1 (0.3 – 4.0) | 0.39 | 1.4 (0.5 – 3.7) | 0.54 | 1.2 (0.4 – 3.4) | 0.78 |
| **Time since positive PCR** | | | | | | | | | | |
| - **<30 days** | 1 | | 1 | | 1 | | 1 | | 1 | |
| - **>30 days** | 2.0 (0.8 – 5.1) | 0.14 | 0.2 (0.1 – 0.8) | 0.03 | 1.1 (0.3 – 3.7) | 0.94 | 0.7 (0.3 – 1.9) | 0.71 | 1.2 (0.5 – 3.0) | 0.71 |

Irwin N, Murray L, Ozynski B, Richards GA, Paget G, Venturas J, Kalla I, Diana N, Mahomed A, Zamparini J. **Age significantly influences the sensitivity of SARS-CoV-2 rapid antibody assays**
